# Early detection of Alzheimer’s disease pathophysiology using 3D virtual reality navigation: a correlational study with genetic and plasma biomarkers

**DOI:** 10.1101/2024.05.01.24306489

**Authors:** Sayuri Shima, Reiko Ohdake, Yasuaki Mizutani, Harutsugu Tatebe, Riki Koike, Atsushi Kasai, Epifanio Bagarinao, Akihiro Ueda, Mizuki Ito, Junichi Hata, Shinsuke Ishigaki, Takahiko Tokuda, Akihiko Takashima, Hirohisa Watanabe

## Abstract

**BACKGROUND:** We investigated the association between path-integration (PI) errors related to entorhinal cortex function detectable using a 3D virtual reality (VR) navigation system and various biomarkers to explore its potential as an early AD indicator.

**METHODS:** The PI capabilities of 111 healthy adults were assessed using a head-mounted 3D VR system. Demographic and cognitive assessments, AD-related plasma biomarkers, and apolipoprotein E genotypes were also evaluated. Predictive factors for PI errors were identified using multivariate linear regression, logistic regression, and random forest.

**RESULTS:** PI errors positively correlated with age, plasma levels of glial fibrillary acidic protein [GFAP], neurofilament light, and p-tau181. Multivariate analysis identified plasma GFAP and p-tau181 levels as significant predictors. Random forest analysis and receiver operating characteristic curves underscored plasma p-tau181 levels as the most substantial predictor.

**DISCUSSION:** PI errors, particularly in conjunction with plasma p-tau181 levels, could reflect early AD pathophysiology, highlighting their potential as early biomarkers.

## 1. BACKGROUND

In Alzheimer’s disease (AD), the entorhinal cortex (EC) is the initial site for the development of neurofibrillary tangles (NFTs), which marks the onset of this neurodegenerative condition<u>[1,2]</u>. These tangles progressively extend from the EC through the limbic cortex to the neocortex, a process that is correlated with the severity of cognitive decline<u>[3]</u>. Although the presence of NFTs in the EC does not directly correlate with dementia symptoms, their expansion into limbic and neocortical areas is commonly associated with mild cognitive impairment (MCI) and overt dementia, respectively<u>[4–6]</u>. Thus, the stages of NFT accumulation offer a pathological timeline of clinical brain aging, with changes in the EC serving as an early indicator of potential progression from a pre-clinical state to clinical deterioration.

The importance of the EC extends beyond its vulnerability to NFTs. The EC contains grid cells that underpin the spatial mapping and navigation capabilities of the brain<u>[7]</u>. These cells are involved in specific periodic self-location representations, which are crucial for path integration (PI). The grid cell network in the EC calculates one’s current position by continuously updating the orientation and movement over time using self-motion cues from visual, vestibular, and proprioceptive sources, independent of external cues such as landmarks<u>[8]</u>. Interestingly, disruptions in grid cell functioning<u>[9]</u> as well as inhibition of EC cells<u>[10]</u> impair PI, and our previous research utilizing P301S mutant tau-overexpressing mice showed that tau pathology in the EC can directly impact PI<u>[10]</u>.

Capitalizing on these insights, several studies introduced a novel approach to early AD detection using a head-mounted 3D virtual reality (VR) system<u>[10–13]</u>. This immersive tool not only encompasses the user’s field of vision but also accurately simulates head and bodily movements, offering a unique method for assessing EC-dependent PI and hippocampus-dependent spatial memory functions. Our previous findings suggested that the 3D VR paradigm can effectively differentiate between these two types of spatial cognition, revealing that PI errors increase with age and may commence around the age of 50, according to Braak’s classification<u>[10]</u>. This correlation between the age-related decline in PI ability and NFT accumulation in the EC, although not directly measured in our participants, underscores the potential of our VR system as a digital biomarker for the detection of early-stage AD.

Recent research underscores the critical need to develop validated clinical protocols that allow physicians to identify individuals at risk of AD at an ultra-early stage, thereby facilitating a preventative approach<u>[14]</u>. There is a consensus that lifestyle modifications can influence up to 40% of AD risk factors<u>[15]</u>. Given these insights, it is paramount to identify molecular biomarkers to detect early pathological changes before the appearance of clinical symptoms. However, molecular markers are generally expensive require invasive procedures, and do not correlate with neurocognitive function and behavioral tests; therefore, it is important to develop simple noninvasive clinical indicators.

This study aimed to elucidate the relationships between PI assessed using a 3D VR system and aging, AD-related biomarkers, EC thickness, and the presence of the apolipoprotein E (*APOE*) ε4 allele in a cohort of 111 healthy individuals. By leveraging this innovative 3D VR navigation system, we seek to shed light on the pathological progression of AD and advance the development of noninvasive diagnostic tools for early detection. Through this approach, we hope not only to contribute to the understanding of the mechanisms underlying AD but also to facilitate early identification and potential intervention in the disease trajectory.

## 2. METHODS

### 2.1. Participants

Data from healthy volunteers, who participated in our ongoing aging registry study at Fujita Health University, were analyzed. The participants were recruited between September 2021 and June 2023, and 140 healthy adults were initially enrolled. Our study was conducted exclusively in Japan, involving a homogeneous population of Japanese participants. This approach was taken due to the study’s geographical and logistical constraints. We acknowledge the limitations this imposes on the diversity of our sample. Future studies should aim to include a broader demographic to enhance the generalizability of our findings.

We collected demographic data, including education, medical history, medication, drinking and smoking habits, and family history of neurodegenerative diseases. All participants underwent 3 T MRI examination and clinical assessments of general cognitive performance using the Mini-Mental State Examination (MMSE)<u>[16]</u>, Japanese version of Addenbrooke’s Cognitive Examination-Revised (ACE-R)<u>[17]</u>, and Japanese version of the Montreal Cognitive Assessment (MoCA-J)<u>[18]</u>. Mood disturbances were evaluated using the Geriatric Depression Scale-15 (GDS-15).

Of the 140recruited participants, 29 were excluded because of (a) inability to complete the 3D navigation task due to VR-induced sickness (9 participants), (b) cognitively abnormal MMSE scores below 26 or ACE-R total scores below 89 (13 participants), (c) medical history of stroke (2 participants) or neurological or psychiatric disorders (3 participants), and (d) incomplete data (2 participants). The characteristics of the 111 finally included participants aged 22 to 79 years are summarized in Table 1.

**TABLE 1.**
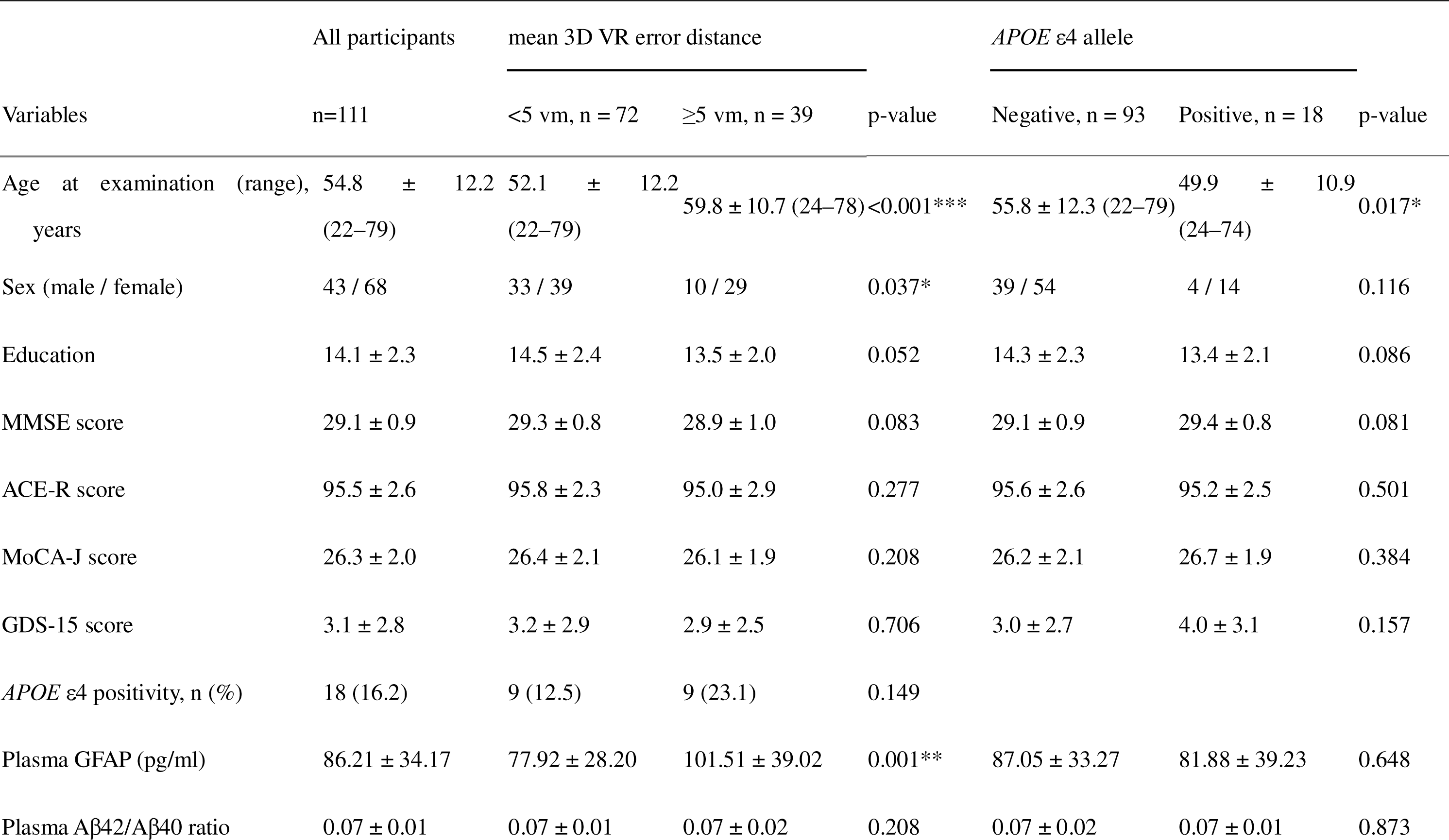
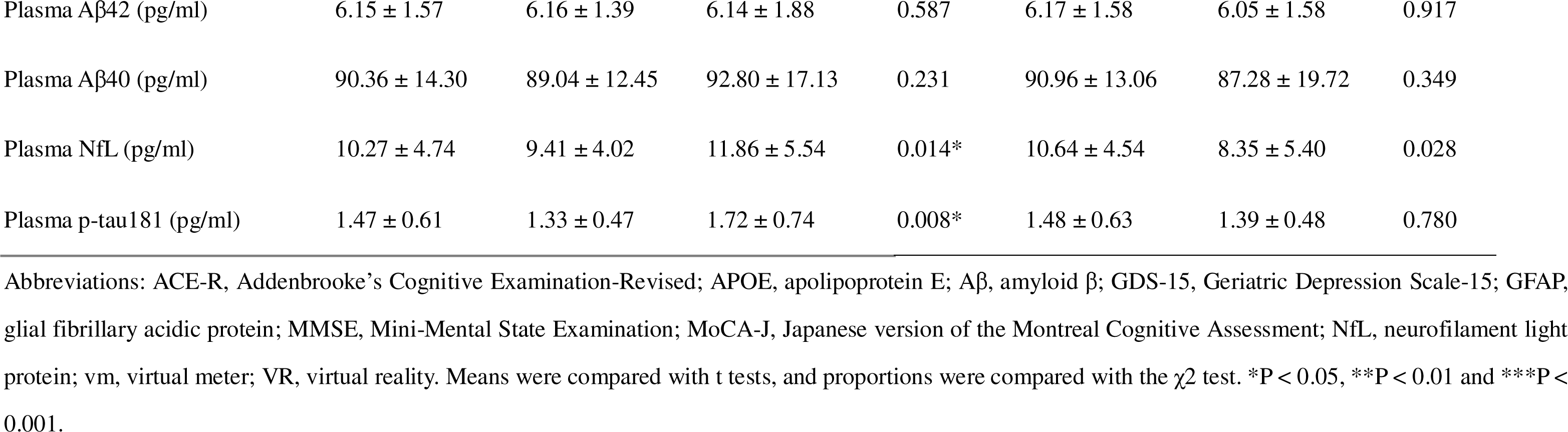

This research was approved by the Ethics Committee at Fujita Health University Hospital (approval number: HM22-407) and conformed to the Ethical Guidelines for Medical and Health Research Involving Human Subjects, as endorsed by the Japanese government. All participants provided written informed consent and retained the option to withdraw from the study at any point prior to joining the study.

### 2.2. 3D VR navigation task

We developed digital biomarkers for the early detection of AD using head-mounted immersive 3D VR devices. These systems are characterized by their capacity to envelop the user’s field of vision and simulate authentic head or body motions<u>[10]</u>. The VR space comprised a 20 virtual meter (vm) diameter arena encircled by walls 3 vm in height. Participants wore 3D VR goggles and controlled their movements using a joystick (Meta Quest 2). Initially, they were permitted to move freely within the virtual arena, including obstacles, to become accustomed to the setup. Forward and backward movements were joystick-controlled, whereas lateral movements required participants to rotate their bodies while seated on a swivel chair. Volunteers with visual impairments or those unable to effectively use a joystick were excluded. Eight participants who had experienced VR sickness were excluded from the analysis. The software and VR goggles described in a previous study<u>[12]</u> were provided by MIG Inc. (Tokyo, Japan; https://www.medicalig.com).

PI performance was assessed in participants wearing the VR goggles who navigated first to a designated Location A (marked by a yellow flag), proceeded then to a designated Location B (marked by a red flag), and finally returned to the starting point (Supplementary Figure S1). During the return to the starting point, all flag markers, including those at Location B, were no longer visible, requiring participants to rely solely on their memory and spatial awareness to navigate back. The distance between the participant’s final position and the actual starting point (error distance) was recorded. The participants completed this test three times, with the average error distance used as the measure of PI ability.

### 2.3. Sample collection and assays of plasma biomarkers

Blood plasma was obtained from all recruited participants by collecting blood samples after a fasting period of more than 6 hours. The samples were centrifuged for 10 min at 1,500 × g. To prevent repeated freeze-thaw cycles, the obtained plasma was aliquoted into 500 µl samples which were promptly frozen and stored at −80°C until analysis. The levels of plasma glial fibrillary acidic protein (GFAP), neurofilament light (NfL) protein, amyloid β (Aβ)40, Aβ42, and p-tau181 were measured using a single-molecule array (Simoa) with the Simoa Human Neurology 4-Plex A Kit and Simoa pTau-181 V2 Advantage Kit (Quanterix, Billerica, MA, USA), following the manufacturer’s instructions. All plasma samples were tested in duplicates.

### 2.4. APOE genotyping

We used the Invader® assay for APOE genotyping, as previously described<u>[19,20]</u>. In brief, the reaction mixture was prepared by combining 1.2 µl of the primary probe/Invader oligo mix with 1.8 µl of Cleavase X Invader core reagent kit (genomic DNA, Hologic) The primary probes/Invader oligo mixture contained 3.5 µmol/l of each wild-type and mutant primary probe and 0.35 µmol/l Invader oligo in 10 mmol/l MOPS. Three microliters of the reaction mixture were added to each well of a 384-well plate. To this, 3 µl of 5 fmol/l synthetic target oligonucleotides (positive control), 10 mg/ml yeast tRNA (for non-target blanks), and genomic DNA (30 ng/µl) were added and denatured by heating at 95°C for 5 min.

After the addition of 6 µl of mineral oil (Sigma, St. Louis, MO) to all reaction wells to prevent evaporation, the plate was incubated at a constant temperature of 63°C for 4 h in a Block Incubator (BI-536; ASTEC, Tokyo, Japan) and then maintained at 15°C until fluorescence measurements were taken. Fluorescence intensity was measured using a fluorescence microtiter plate reader (Infinite 200 PRO F Nano+; TECAN, Kanagawa, Japan) with excitation at 485 nm/20 nm and emission at 535 nm/25 nm for FAM and excitation at 560 nm/20 nm and emission at 612 nm/10 nm for RED.

Genotyping analysis was performed by calculating the ratio of net fluorescence counts for the wild-type primary probe to that of the mutant primary probe.

### 2.5. MRI data

All participants underwent MRI scanning at Fujita Health University using a 3 T MRI scanner (Canon Medical Systems) with a 32-channel head coil. For each participant, T1-weighted images were acquired using a 3D fast field echo sequence with the following imaging parameters: repetition time = 6.6/2500 ms, echo time = 2.7 ms, flip angle = 8 degrees, field-of-view = 240 × 240 mm2, acquisition/reconstruction matrix = 288 × 288, 230 sagittal slices with 0.8-mm slice thickness, and voxel resolution of 0.8 × 0.8 × 0.8 mm3 with a total acquisition time of 5 min and 5 s.

All T1-weighted images were preprocessed with Freesurfer v7.2 (https://surfer.nmr.mgh.harvard.edu/) using its default *recon-all* pipeline. Regional volume and thickness data were then extracted from the preprocessed images using the Desikan-Killiany atlas<u>[21]</u>, which consists of 34 cortical regions of interest (ROIs) per hemisphere.

### 2.6. Statistical analyses

We conducted various statistical analyses to examine the relationship between PI performance and various demographic, clinical, and biomarker measures. We employed independent samples of the Mann-Whitney U test and the chi-square tests for demographic comparisons between the two independent study groups. Correlation analyses of the extracted volume and thickness data were performed using PI and biomarker data. Pearson’s correlation coefficients and their corresponding p-values were computed using the *corr()* function in MATLAB (R2023a; MathWorks, Natick, Mass, USA). We also examined the association between PI and EC thickness and volume using linear regression, which included age, sex, and estimated total intracranial volume (for volume data only) as covariates. To elucidate the factors that significantly predicted PI errors, we performed multivariate linear regression analyses by implementing the standard least squares method. We evaluated the predictive power of age, GFAP, NfL, p-tau181, Aβ40, Aβ42, the ratio of Aβ42/Aβ40 ratio, and *APOE* ε4 allele status.

Logistic regression was performed to determine the odds ratios for PI errors exceeding 5 vm based on the same set of predictors. The Wald chi-square test was used to determine the significance of each predictor within the model. The performance of the random forest model was evaluated using measures such as the Mean Decrease in Accuracy and the Mean Decrease in Gini, which indicate the importance of each predictor variable in the model. The accuracy of the final model was assessed using the area under the curve (AUC), and the sensitivity and specificity were calculated at the optimal cut-off point determined by Youden’s index.

Receiver operating characteristic (ROC) curve analysis was used to determine the optimal plasma p-tau181 level cut-off for predicting significant PI errors. We set the cutoff at 2.2 pg/ml, which shows high accuracy in discriminating AD from other disease groups and healthy people (in-house cut-off value of Institute for Quantum Medical Science). The AUC, standard error of the AUC, and 95% confidence interval were computed to assess the accuracy of the p-tau181 level as a predictive biomarker. The sensitivity and specificity of these cut-off values were also calculated to understand the practical applicability of our findings in a clinical setting.

All statistical analyses were performed using JMP software version 16 (SAS Institute, Cary, NC, USA), and an alpha level of 0.05 was set to determine statistical significance.

## 3. RESULTS

### 3.1. Demographics

Table 1 presents the demographic characteristics of the study groups. The mean age at examination was 54.8 ± 12.2 (22–79) years in the entire study population, and women comprised 61.3%. The 39 participants (35.1%) with ≥5 vm error distance had a higher age at examination, a higher percentage of female participants, and elevated plasma levels of GFAP, NfL, and p-tau181 compared to those with <5 vm error distance. General cognitive performance assessed by the MMSE, ACE-R, and MoCA-J was not significantly different between these two groups. *APOE* ε4 was positive in 18 participants (16.2%) of the entire study population. Participants with *APOE* ε4 positivity were younger than those without.

### 3.2 Relationship between PI errors and age and plasma AD-related biomarkers

PI errors assessed using 3D VR goggles showed significant positive correlations with age (r = 0.304, p = 0.0012) and plasma levels of GFAP (r = 0.334, p = 0.0003), p-tau181 (r = 0.327, p = 0.0005), and NfL (r = 0.205, p = 0.0310; Figure 1). Notably, no significant correlations were found between PI errors and plasma levels of Aβ40 and Aβ42, as well as the Aβ42/Aβ40 ratio. These findings are visually supported by a comprehensive heatmap that details the strength and direction of relationships between measured biomarkers and PI performance (Supplementary Figure S2).

**FIGURE 1.**
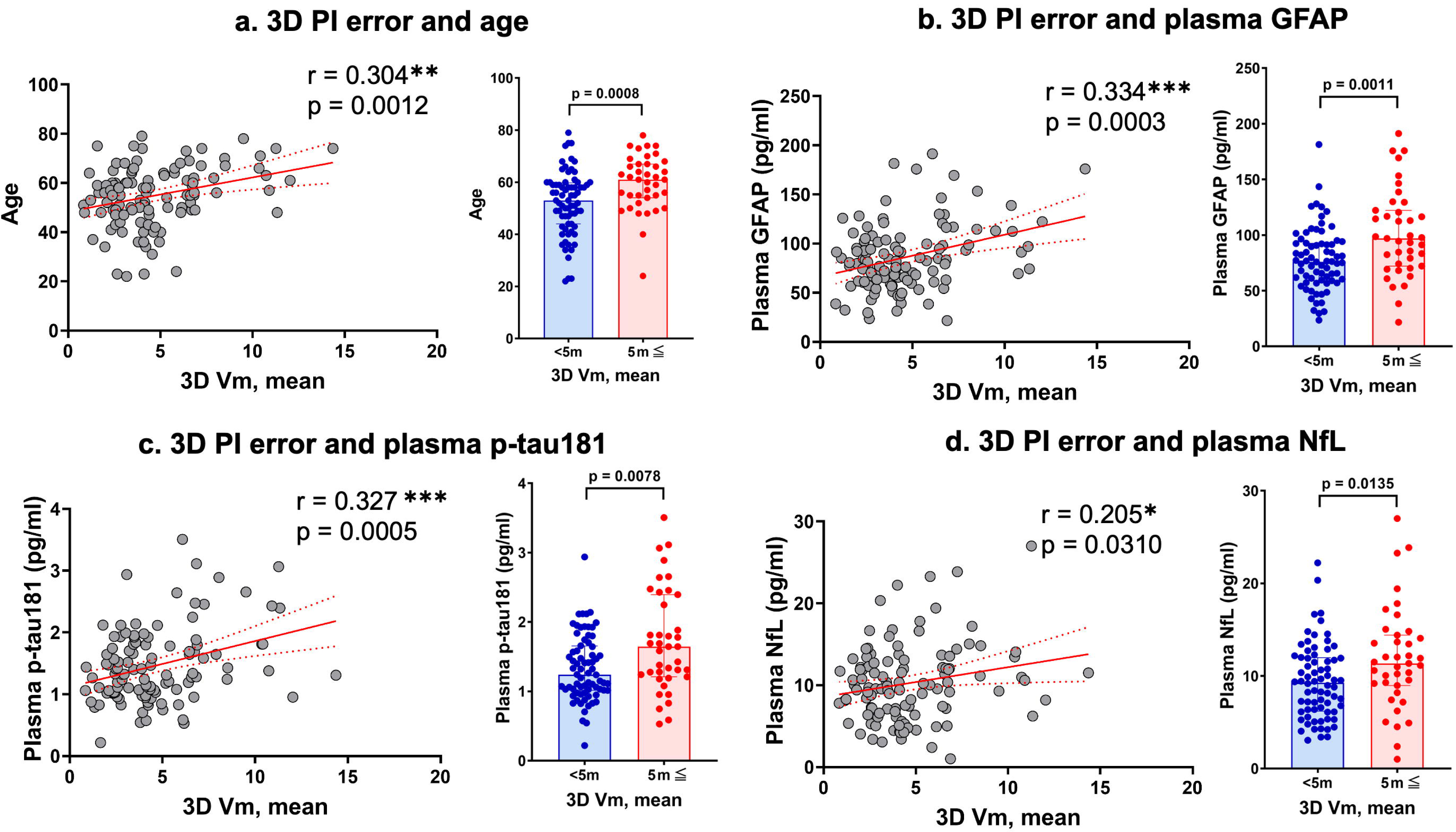
Correlations between 3D VR path integration errors and various markers. Panel a shows the relationship between mean error distance in the 3D VR navigation task and participant age. Panels b, c, and d depict the correlations of mean error distance with plasma levels of GFAP, p-tau181, and NfL, respectively. The scatter plots are complemented by bar graphs comparing groups with mean error distances of <5 vm and ≥5 vm. Solid red lines represent regression lines, and red dashed lines show 95% confidence intervals. Significance levels are marked as *p<0.05, **p<0.01, and ***p<0.001. Abbreviations: GFAP, glial fibrillary acidic protein; NfL, neurofilament light; vm, virtual meter; PI, path integration; VR, virtual reality.

Multivariate analysis utilizing standard least squares revealed that plasma GFAP and p-tau181 levels were significantly correlated with PI errors (Table 2). In the logistic regression analysis, both plasma p-tau181 levels and the presence of the *APOE* ε4 allele were significantly associated with increased PI errors (Table 3).

**TABLE 2.**

| Variable | Coefficient | Std. error | t-value | P-value | 95% CI |
| --- | --- | --- | --- | --- | --- |
| Intercept | 0.102767 | 6.660252 | 0.02 | 0.9877 | (-13.10781, 13.313346) |
| Age | 0.057416 | 0.029592 | 1.94 | 0.0551 | (-0.001279, 0.1161117) |
| Plasma Aβ40 | -0.007057 | 0.077906 | -0.09 | 0.928 | (-0.161582, 0.1474683) |
| Plasma Aβ42 | 0.035813 | 1.165466 | 0.03 | 0.9755 | (-2.275884, 2.3475103) |
| Plasma Aβ42/Aβ40 ratio | -0.472902 | 100.471100 | 0.00 | 0.9963 | (-199.7569, 198.81112) |
| Plasma GFAP | 0.019661 | 0.009108 | 2.16 | 0.0332* | (0.0015949, 0.0377279) |
| Plasma NfL | -0.140732 | 0.084714 | -1.66 | 0.0997 | (-0.308763, 0.0272984) |
| Plasma p-tau181 | 1.176500 | 0.464560 | 2.53 | 0.0128* | (0.2550471, 2.0979536) |
| *APOE* ε4 positivity | -0.160592 | 0.328495 | -0.49 | 0.626 | (-0.812161, 0.4909761) |

Abbreviations: APOE, apolipoprotein E; Aβ , amyloid $\beta$ ; CI, confidence interval; GFAP, glial fibrillary acidic protein; NfL, neurofilament light protein. \*P < 0.05.

**TABLE 3.**

| Predictor | Coefficient | Std. error | Wald chi-square | P-value | (95% CI) |
| --- | --- | --- | --- | --- | --- |
| Interception | -0.7562064 | 6.2110111 | 0.01 | 0.9031 | (-12.906729, 12.4260344) |
| Age | 0.06140072 | 0.0322009 | 3.64 | 0.0565 | (0.00123923, 0.12854751) |
| Plasma Aβ40 | -0.0579735 | 0.0726146 | 0.64 | 0.4247 | (-0.213181, 0.08301344) |
| Plasma Aβ42 | 0.77163763 | 1.1166312 | 0.48 | 0.4895 | (-1.3612706, 3.15317088) |
| Plasma Aβ42/Aβ40 ratio | -65.460018 | 96.369905 | 0.46 | 0.497 | (-273.39975, 116.672357) |
| Plasma GFAP | 0.01464229 | 0.0084491 | 3 | 0.0831 | (-0.001549, 0.03193665) |
| Plasma NfL | -0.0577238 | 0.0777763 | 0.55 | 0.458 | (-0.2135507, 0.09552579) |
| Plasma p-tau181 | 0.89940611 | 0.4408718 | 4.16 | 0.0413* | (0.05817076, 1.8023939) |
| *APOE* ε4 positivity | -0.650281 | 0.3121227 | 4.34 | 0.0372* | (-1.2847926, -0.0475706) |

Abbreviations: APOE, apolipoprotein E; Aβ , amyloid $\beta$ ; CI, confidence interval; GFAP, glial fibrillary acidic protein; NfL, neurofilament light protein. \*P < 0.05.

Using random forest analysis, a comprehensive evaluation was conducted to identify the predictors of PI errors. This machine learning approach revealed that among the array of tested variables, the plasma p-tau181 levels emerged as the most significant predictor, which is consistent with our logistic regression findings. Further analysis using the random forest model provided a hierarchy of variable importance, with p-tau181 at the forefront, followed by the other plasma biomarkers and genetic factors. Although the *APOE* ε4 allele was recognized as a factor in the logistic regression analysis, its predictive power was comparatively lower in the random forest model (Supplementary Figure S3).

### 3.3. ROC curves for plasma p-tau181 levels

The ROC curve for detecting plasma p-tau181 levels ≥ 2.2 presented an area under the curve (AUC) of 0.8577, suggesting a high degree of accuracy in differentiating individuals based on PI errors. The standard error for the AUC was 0.05241, and the 95% confidence interval ranged from 0.7550 to 0.9605, indicating strong model performance. The optimal cut-off value for 3PI errors was determined to be 5.77746 vm, achieving a sensitivity of 91.7% and specificity of 77.8%. The statistical significance of this model was supported by a p-value below 0.0001 (Figure 2).

**FIGURE 2.**
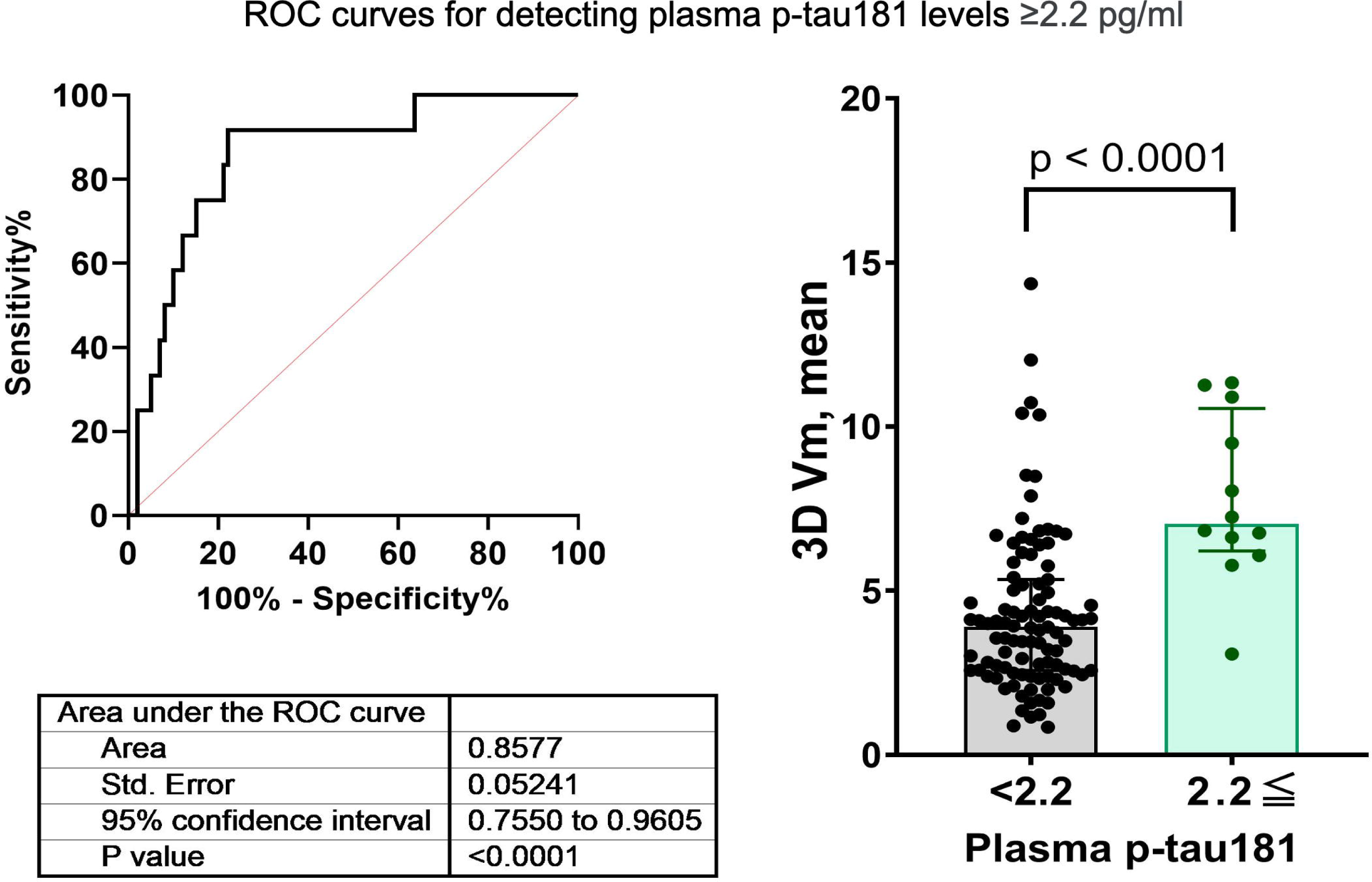
ROC curves assessing the diagnostic performance of plasma p-tau181 levels for detecting path integration errors. Panels a and b illustrate the ROC curves for a p-tau181 level cutoff of 2.3 and 1.98 pg/ml, respectively, along with their respective AUCs, standard errors, confidence intervals, and p-values. Abbreviations: AUC, area under the curve; ROC, receiver operating characteristic.

### 3.4. Associations with EC thickness

The thickness of the left EC was negatively correlated with age (r = -0.4310, p = 2.34 × 10-6) and mean PI error (r = -0.2129, p = 0.0249) (Supplementary Figure S4), as well as with several biomarkers, including Aβ40 (r = -0.2011, p = 0.0343), GFAP (r = -0.1971, p = 0.0381), NfL (r = -0.2328, p = 0.0139), and p-tau181 (r = -0.2373, p = 0.0122) levels) Likewise, the thickness of the right EC was negatively correlated with age (r = -0.3628, p = 9.11 × 10-5), Aβ40 level (r = -0.2242, p = 0.0180), and NfL level (r = -0.2560, p = 0.0067). In the linear regression, which included age and sex as covariates, the association between the thickness of the right and left EC and PI error was not significant (Supplementary Figure S5). No significant associations were observed between EC volume and these measures.

## 4. DISCUSSION

Our analysis revealed that PI errors, assessed using the 3D VR navigation system, were significantly correlated with older age and elevated plasma levels of AD-related biomarkers including GFAP, p-tau181, and NfL. Results of multivariate analysis suggested that plasma GFAP level, APOE ε4 positivity, and especially plasma p-tau181 level are more significant predictors of PI performance than age. Further analysis using the random forest model provided a hierarchy of variable importance, with plasma p-tau181 level at the forefront. These findings indicate that the observed PI errors are not solely a consequence of aging but may also reflect the underlying AD pathology. Our results highlight the utility of the 3D VR navigation system in capturing these deficits, suggesting its effectiveness as an early surrogate marker for pathophysiological changes that are indicative of AD.

Recent advancements have highlighted the potential of plasma p-tau181 levels as a noninvasive biomarker for AD<u>[22,23]</u>, although it is unclear whether the observed p-tau181 levels reflect tau deposition in the EC because plasma p-tau181 levels are not only related to tau PET but also to amyloid PET. This biomarker not only predicts disease progression but also acts as an early pathological marker, with elevated levels correlating with the initial neuropathological changes<u>[24]</u>. The current study found a significant relationship between PI errors and plasma p-tau181 levels, suggesting that these abnormalities may reflect the progression of AD pathology. Moreover, ROC analyses showed that PI errors can predict increased plasma levels of p-tau181. A combination of AD-related biomarkers and PI errors assessed using the 3D VR navigation system may distinguish whether these decreases are attributable to normal aging or early pathological changes specific to AD, thereby enhancing our ability to detect and monitor AD at an early stage.

Not only p-tau181 but also plasma phospho-tau, such as p-tau217 and p-tau231, have become available<u>[25]</u> and can predict amyloid PET and tau PET stages<u>[26]</u>. 18F-MK-6240 revealed topographical patterns consistent with Braak stage<u>[27]</u> and even captured the Braak stages I and II<u>[28]</u>. Therefore, further studies are needed to explore the relationship between plasma p-tau217 or p-tau231 levels, tau deposition in the EC indicated by MK-6240 PET, and abnormalities in PI capabilities.

This is the first study to show that PI errors can correlate with AD-related biomarkers particularly p-tau181 before the onset of cognitive decline. Recently, Newton et al. reported that entorhinal-based PI selectively predicts the midlife risk of AD in asymptomatic adults, linking it to early neurodegenerative changes before other cognitive declines are observable<u>[29]</u>. Our results also confirmed that the presence of the APOE ε4 allele is significantly associated with increased PI errors<u>[13,30,31]</u>. These results support the idea that immersive VR to measure PI can effectively reveal early-stage AD-related changes. Moreover, the specific impairment in PI abilities noted in our findings, independent of traditional cognitive decline, supports the notion that PI might be among the first detectable signs of cognitive alteration in AD progression, and can facilitate early, effective therapeutic interventions.

The thickness of the EC in univariate analyses appeared to be negatively correlated with age and mean PI errors, as well as with several biomarkers, suggesting that thinner cortical regions might be associated with greater navigational difficulties and cognitive markers of AD. However, these associations were not statistically significant after adjusting for age and other confounding variables. This finding is consistent with a previous report that investigated the VR PI ability in asymptomatic middled-aged adults stratified by hereditary and physiological AD risk factors<u>[29]</u>. Interestingly, EC thickness was correlated with PI ability in people with MCI<u>[12]</u>. These findings emphasize the functional impairment of the EC even before significant structural changes become detectable, indicating that EC thickness changes may parallel the progression of navigational and cognitive decline in AD<u>[4]</u>; they may be independent predictors when controlling for age.

PI capabilities were also correlated with plasma GFAP levels but not with the plasma Aβ42/Aβ40 ratio, which has lower sensitivity than the corresponding ratio in the cerebrospinal fluid and in amyloid PET data<u>[32]</u>. GFAP, an astrocytic cytoskeletal protein with increased blood levels in individuals with AD and MCI, is especially upregulated around Aβ plaques and is correlated with tau accumulation, indicating its involvement in neuroinflammatory responses and astrocytic reactivity<u>[33]</u>. The lack of correlation between the plasma Aβ42/Aβ40 ratio and PI ability might be caused not only by the low sensitivity of the plasma Aβ42/Aβ40 ratio. The presented correlation between plasma GFAP and PI performance, independent of the plasma Aβ42/Aβ40 ratio, suggests GFAP’s potential as a reactive marker to neuroinflammation. It is essential to clarify that the lack of association with the Aβ42/Aβ40 ratio could stem from multiple factors. These include the inherently lower sensitivity of plasma Aβ measures compared to cerebrospinal fluid or PET imaging, the specificity of GFAP as a neuroinflammatory marker, the timing of its elevation in the disease course, and the possible disconnect between soluble biomarkers and actual plaque deposition. Further research is needed to elucidate the distinct roles of these biomarkers in AD pathogenesis and their interrelationships.

PI is primarily associated with egocentric navigation, which relies on an individual’s internal perspective to navigate and maintain orientation based on their own movements and sensations such as acceleration and direction changes<u>[34]</u>. It involves continuously updating one’s position relative to the starting point without requiring external landmarks. PI involves integrating the continuous signals related to the individual’s movements, such as the direction turned, the number of steps taken, and the speed and angle of movements, integrating self-motion cues provided by the visual, proprioceptive, and vestibular systems<u>[8]</u>. Thus, age-related decline in sensory function may also be associated with deficits in egocentric navigation<u>[35]</u>. While our study did not directly measure primary sensory functions, such as optic flow or vestibular responses, the strong association between p-tau181 level and navigation deficits underscores the potential role of early neurodegenerative processes affecting these sensory pathways. Our previous study demonstrated the importance of maintaining sensorimotor, auditory, and visual networks in preserving cognitive function in older adults<u>[36]</u>. Furthermore, the known association between hearing loss as a risk factor for AD<u>[15]</u> and reports suggesting that cataract extraction may reduce the risk of AD<u>[37]</u> underscores the critical role of primary sensory information in cognitive health. These findings suggest that deficits in primary sensory inputs might not only reflect sensory deterioration but may also have profound effects on EC function. Future studies should aim to directly assess these sensory functions and determine their specific contribution to the navigation deficits observed in early AD and whether they may cause AD pathology in the EC.

### 4.1. Limitation

Our study has several limitations that warrant consideration. First, while our 3D VR navigation system has demonstrated potential for detecting early PI deficits, it does not evaluate fundamental sensory information, such as visual flow, vestibular function, and proprioception, which are integrated within the EC and decline with aging. This limitation suggests that our findings may reflect not only early markers of AD but also normal age-related changes, making it challenging to distinguish between the two. Second, the absence of detectable changes in AD-related plasma biomarkers in participants with PI errors highlights the need for a comprehensive approach that includes a broader range of diagnostic modalities to fully capture the early stages of AD pathophysiology. Furthermore, the imbalance in sample size for participants with the APOE ε4 allele might affect the generalizability of our results. Future studies incorporating these sensory evaluations and involving larger and more diverse cohorts are essential to validate our findings and refine the utility of VR systems in early AD detection. While our study provides valuable insights into the relationship between path integration errors and plasma p-tau181 levels among cognitively normal Japanese individuals, the lack of ethnic and racial diversity may limit the applicability of our results to other populations. Furthermore, due to the exploratory nature of our research and the unknown prevalence of AD biomarker abnormalities in cognitively normal populations, a precise sample size calculation was not feasible. Future research should consider these factors to strengthen the study design and potential findings.

### 4.2. Conclusions

Our study substantiates the efficacy of the 3D VR navigation system in capturing PI errors that may reflect early AD pathophysiology, particularly in terms of plasma p-tau181 levels. These findings advance the application of digital biomarkers for AD and advocate for the broader adoption of similar non-invasive diagnostic tools in clinical settings. While our results are promising, we acknowledge the need for larger studies to confirm these associations and explore the influence of other potential biomarkers. Ultimately, integrating such innovative diagnostic approaches may revolutionize the early detection and monitoring of AD, paving the way for timely therapeutic interventions and improved patient outcomes.

## Supporting information

Supplementary flie1-5

## Data Availability

All data produced in the present work are contained in the manuscript

## Abbreviations

ACE-R: Addenbrooke’s Cognitive Examination-Revised
AD: Alzheimer’s disease
APOE: apolipoprotein E
AUC: area under the curve
Aβ: amyloid β
EC: entorhinal cortex
GDS-15: Geriatric Depression Scale-15
GFAP: glial fibrillary acidic protein
MCI: mild cognitive impairment
MMSE: Mini-Mental State Examination
MoCA-J: Japanese version of the Montreal Cognitive Assessment
NfL: neurofilament light
NFTs: neurofibrillary tangles
PI: path integration
ROC: receiver operating characteristic
vm: virtual meter
VR: virtual reality

## ACKNOWLEDGMENTS

MIG Inc. provided equipment and helped with measurements using the 3D VR goggles. We would like to thank Editage (www.editage.jp) for English language editing.

---

**Figure S1** Schematic representation of the 3D VR navigation task. Path integration was measured in VR goggle-wearing participants going to Location A (indicated by a yellow flag), then to Location B (indicated by a red flag), and finally returning to the starting point. The error distance between the participant’s final location (x) and the starting point was determined. Abbreviation: VR, virtual reality.

**Figure S2** Heatmap depicting the correlation matrix of path integration errors measured by 3D VR navigation (3D VR, Vm) and various plasma biomarkers. The matrix presents Pearson’s correlation coefficients between variables including plasma Aβ40, Aβ42, Aβ42/Aβ40 ratio, GFAP, NfL, and p-tau181. Positive correlations are indicated in red and negative correlations in blue, with the color intensity representing the correlation strength. Significant correlations are denoted as *p<0.05, **p<0.01, and ***p<0.001. Abbreviations: Aβ, amyloid β; GFAP, glial fibrillary acidic protein; NfL, neurofilament light protein; vm, virtual meter; VR, virtual reality.

**Figure S3** Result of random forest analysis. The application of a machine learning method disclosed that, from the pool of variables examined, the levels of plasma p-tau181 stood out as the principal forecast element, aligning with the outcomes of our logistic regression analysis. Subsequent evaluations employing the random forest model delineated a ranked order of influence among the variables, positioning p-tau181 at the top, with additional plasma biomarkers and genetic determinants trailing behind.

**Figure S4** Pearson’s correlation analysis of the entorhinal cortex thickness and volume versus the mean error distance in the 3D VR navigation task. The left entorhinal cortex is shown in the left panels, and the right entorhinal cortex is displayed in the right panels, both with corresponding p-values. Solid black lines represent regression fits, and dashed lines represent 95% confidence intervals. Abbreviation: VR, virtual reality.

**Figure S5** Linear regression analysis of the entorhinal cortex thickness and volume against the mean error distance in the 3D VR navigation task. The left entorhinal cortex is presented in the left panels, and the right entorhinal cortex is shown in the right panels. Statistical significance is denoted where applicable. Solid black lines represent regression fits, and dashed lines represent 95% confidence intervals. Abbreviation: VR, virtual reality.

## References

[1] Braak H, Braak E. Staging of Alzheimer’s disease-related neurofibrillary changes. Neurobiol Aging 1995;16:271–8; discussion 278#x2013;84.

[2] Braak H, Braak E. Evolution of the neuropathology of Alzheimer’s disease. Acta Neurol Scand Suppl 1996;165:3–12.

[3] Jellinger K. Clinicopathological analysis of dementia disorders in the elderly--an update. J Alzheimers Dis 2006;9:61–70.

[4] Rodrigue KM, Raz N. Shrinkage of the entorhinal cortex over five years predicts memory performance in healthy adults. J Neurosci 2004;24:956–63.

[5] Jack CR Jr, Shiung MM, Gunter JL, O’Brien PC, Weigand SD, Knopman DS, et al. Comparison of different MRI brain atrophy rate measures with clinical disease progression in AD. Neurology 2004;62:591–600.

[6] Thaker AA, Weinberg BD, Dillon WP, Hess CP, Cabral HJ, Fleischman DA, et al. Entorhinal Cortex: Antemortem Cortical Thickness and Postmortem Neurofibrillary Tangles and Amyloid Pathology. AJNR Am J Neuroradiol 2017;38:961–5.

[7] Hafting T, Fyhn M, Molden S, Moser M-B, Moser EI. Microstructure of a spatial map in the entorhinal cortex. Nature 2005;436:801–6.

[8] Segen V, Ying J, Morgan E, Brandon M, Wolbers T. Path integration in normal aging and Alzheimer’s disease. Trends Cogn Sci 2022;26:142–58.

[9] Gil M, Ancau M, Schlesiger MI, Neitz A, Allen K, De Marco RJ, et al. Impaired path integration in mice with disrupted grid cell firing. Nat Neurosci 2018;21:81–91.

[10] Koike R, Soeda Y, Kasai A, Fujioka Y, Ishigaki S, Yamanaka A, et al. Path integration deficits are associated with phosphorylated tau accumulation in the entorhinal cortex. Brain Commun 2024;6:fcad359.

[11] Stangl M, Achtzehn J, Huber K, Dietrich C, Tempelmann C, Wolbers T. Compromised Grid-Cell-like Representations in Old Age as a Key Mechanism to Explain Age-Related Navigational Deficits. Curr Biol 2018;28:1108–15.e6.

[12] Howett D, Castegnaro A, Krzywicka K, Hagman J, Marchment D, Henson R, et al. Differentiation of mild cognitive impairment using an entorhinal cortex-based test of virtual reality navigation. Brain 2019;142:1751–66.

[13] Kunz L, Schröder TN, Lee H, Montag C, Lachmann B, Sariyska R, et al. Reduced grid-cell–like representations in adults at genetic risk for Alzheimer’s disease. Science 2015;350:430–3.

[14] González-Madrid A, Calfío C, González A, Lüttges V, Maccioni RB. Toward Prevention and Reduction of Alzheimer’s Disease. J Alzheimers Dis 2023;96:439–57.

[15] Livingston G, Sommerlad A, Orgeta V, Costafreda SG, Huntley J, Ames D, et al. Dementia prevention, intervention, and care. Lancet 2017;390:2673–734.

[16] Folstein M, Folstein M, Folstein S, Folstein S, McHugh P, McHugh P. “Mini-mental state”: A practical method for grading the cognitive state of patients for the clinician. J Psychiatr Res 1975;12:189–98.

[17] Mioshi E, Dawson K, Mitchell J, Arnold R, Hodges JR. The Addenbrooke’s Cognitive Examination Revised (ACE-R): a brief cognitive test battery for dementia screening. Int J Geriatr Psychiatry 2006;21:1078–85.

[18] Nasreddine ZS, Phillips NA, Bédirian V, Charbonneau S, Whitehead V, Collin I, et al. The Montreal Cognitive Assessment, MoCA: a brief screening tool for mild cognitive impairment. J Am Geriatr Soc 2005;53:695–9.

[19] Arai H, Yamamoto A, Matsuzawa Y, Saito Y, Yamada N, Oikawa S, et al. Polymorphisms of apolipoprotein e and methylenetetrahydrofolate reductase in the Japanese population. J Atheroscler Thromb 2007;14:167–71.

[20] Nagano M, Yamashita S, Hirano K-I, Ito M, Maruyama T, Ishihara M, et al. Two novel missense mutations in the CETP gene in Japanese hyperalphalipoproteinemic subjects: High-throughput assay by Invader® assay. J Lipid Res 2002;43:1011–8.

[21] Desikan RS, Ségonne F, Fischl B, Quinn BT, Dickerson BC, Blacker D, et al. An automated labeling system for subdividing the human cerebral cortex on MRI scans into gyral based regions of interest. Neuroimage 2006;31:968–80.

[22] Janelidze S, Mattsson N, Palmqvist S, Smith R, Beach TG, Serrano GE, et al. Plasma P-tau181 in Alzheimer’s disease: relationship to other biomarkers, differential diagnosis, neuropathology and longitudinal progression to Alzheimer’s dementia. Nat Med 2020;26:379–86.

[23] Palmqvist S, Tideman P, Cullen N, Zetterberg H, Blennow K, Alzheimer’s Disease Neuroimaging Initiative, et al. Prediction of future Alzheimer’s disease dementia using plasma phospho-tau combined with other accessible measures. Nat Med 2021;27:1034–42.

[24] Smirnov DS, Ashton NJ, Blennow K, Zetterberg H, Simrén J, Lantero-Rodriguez J, et al. Plasma biomarkers for Alzheimer’s Disease in relation to neuropathology and cognitive change. Acta Neuropathol 2022;143:487–503.

[25] Barthélemy NR, Salvadó G, Schindler SE, He Y, Janelidze S, Collij LE, et al. Highly accurate blood test for Alzheimer’s disease is similar or superior to clinical cerebrospinal fluid tests. Nat Med 2024. 10.1038/s41591-024-02869-z.

[26] Jack CR, Wiste HJ, Algeciras-Schimnich A, Figdore DJ, Schwarz CG, Lowe VJ, et al. Predicting amyloid PET and tau PET stages with plasma biomarkers. Brain 2023;146:2029–44.

[27] Pascoal TA, Therriault J, Benedet AL, Savard M, Lussier FZ, Chamoun M, et al. 18F-MK-6240 PET for early and late detection of neurofibrillary tangles. Brain 2020;143:2818–30.

[28] Krishnadas N, Doré V, Robertson JS, Ward L, Fowler C, Masters CL, et al. Rates of regional tau accumulation in ageing and across the Alzheimer’s disease continuum: an AIBL 18F-MK6240 PET study. eBioMedicine 2023;88. 10.1016/j.ebiom.2023.104450.

[29] Newton C, Pope M, Rua C, Henson R, Ji Z, Burgess N, et al. Entorhinal-based path integration selectively predicts midlife risk of Alzheimer’s disease. Alzheimers Dement 2024. 10.1002/alz.13733.

[30] Bierbrauer A, Kunz L, Gomes CA, Luhmann M, Deuker L, Getzmann S, et al. Unmasking selective path integration deficits in Alzheimer’s disease risk carriers. Sci Adv 2020;6:eaba1394.

[31] Colmant L, Bierbrauer A, Bellaali Y, Kunz L, Van Dongen J, Sleegers K, et al. Dissociating effects of aging and genetic risk of sporadic Alzheimer’s disease on path integration. Neurobiol Aging 2023;131:170–81.

[32] Janelidze S, Teunissen CE, Zetterberg H, Allué JA, Sarasa L, Eichenlaub U, et al. Head-to-Head Comparison of 8 Plasma Amyloid-β 42/40 Assays in Alzheimer Disease. JAMA Neurol 2021;78:1375–82.

[33] Kim KY, Shin KY, Chang K-A. GFAP as a Potential Biomarker for Alzheimer’s Disease: A Systematic Review and Meta-Analysis. Cells 2023;12. 10.3390/cells12091309.

[34] Coughlan G, Laczó J, Hort J, Minihane A-M, Hornberger M. Spatial navigation deficits - overlooked cognitive marker for preclinical Alzheimer disease? Nat Rev Neurol 2018;14:496–506.

[35] Xie Y, Bigelow RT, Frankenthaler SF, Studenski SA, Moffat SD, Agrawal Y. Vestibular Loss in Older Adults Is Associated with Impaired Spatial Navigation: Data from the Triangle Completion Task. Front Neurol 2017;8:173.

[36] Bagarinao E, Watanabe H, Maesawa S, Mori D, Hara K, Kawabata K, et al. Reorganization of brain networks and its association with general cognitive performance over the adult lifespan. Sci Rep 2019;9:11352.

[37] Lee CS, Gibbons LE, Lee AY, Yanagihara RT, Blazes MS, Lee ML, et al. Association Between Cataract Extraction and Development of Dementia. JAMA Intern Med 2022;182:134–41

