## Supplementary figures and images for "Early detection of Alzheimer’s disease pathophysiology using 3D virtual reality navigation: a correlational study with genetic and plasma biomarkers"

### Figure S1.tiff

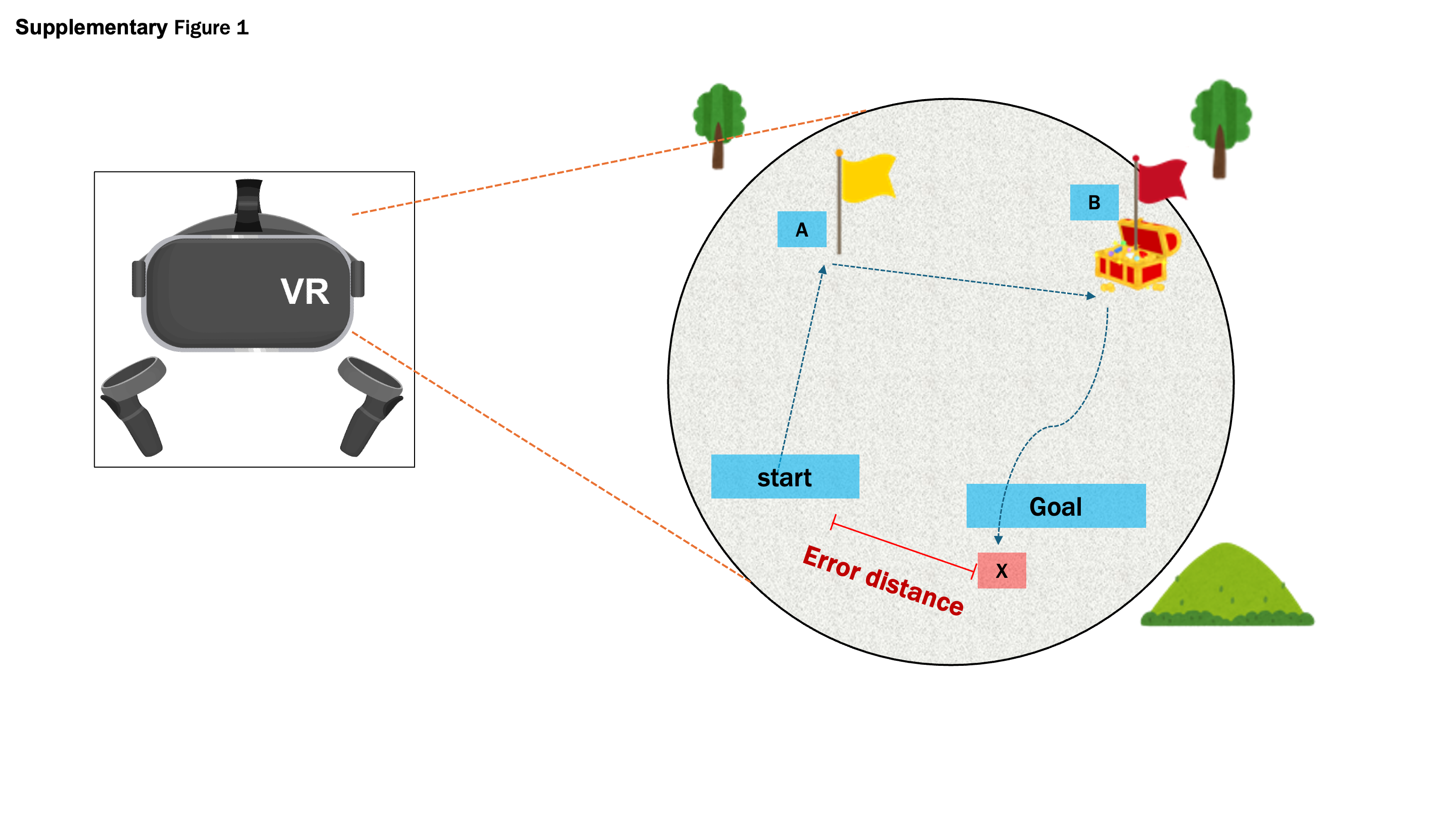

### Figure S2.tiff

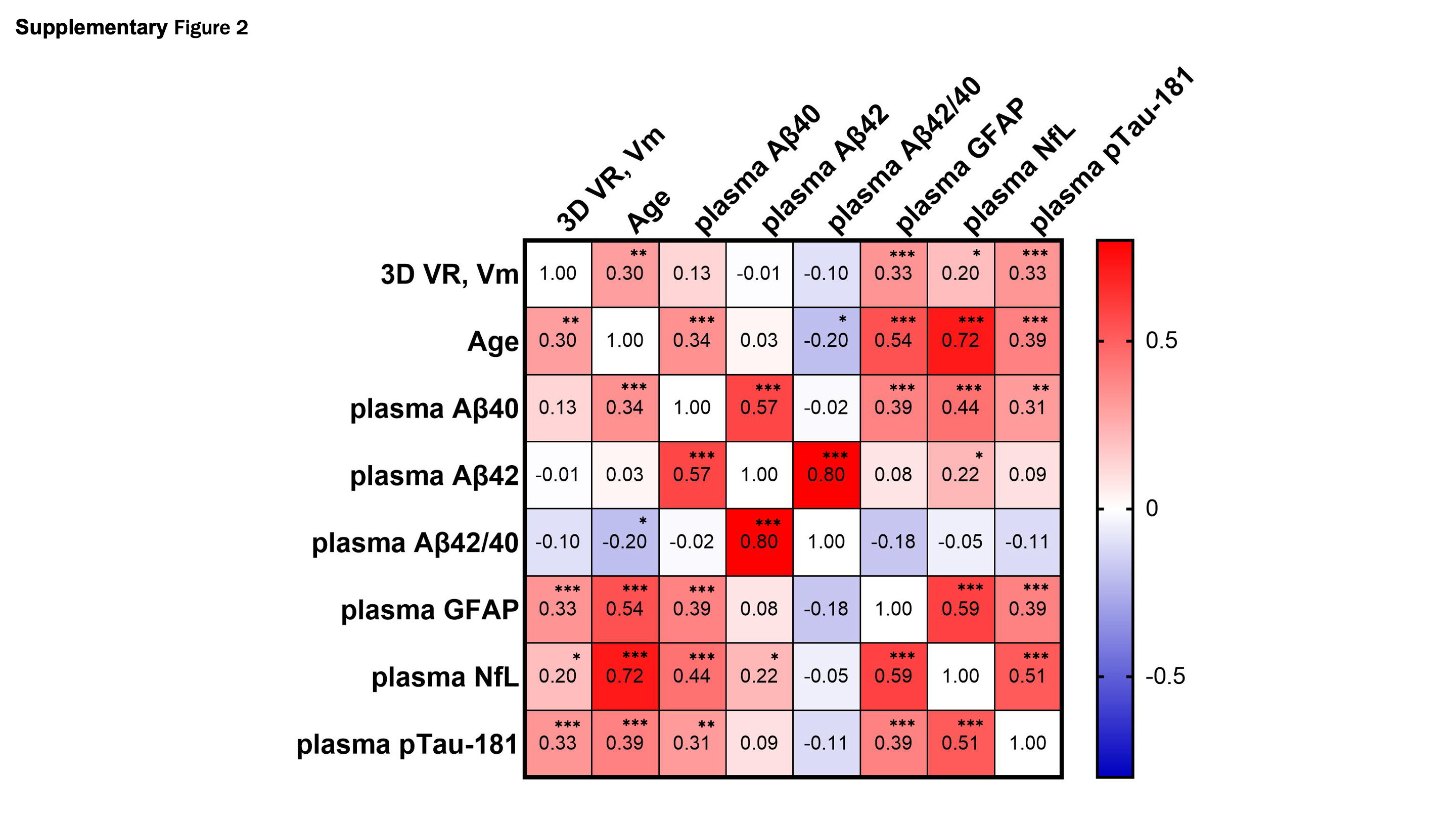

### Figure S3.tiff

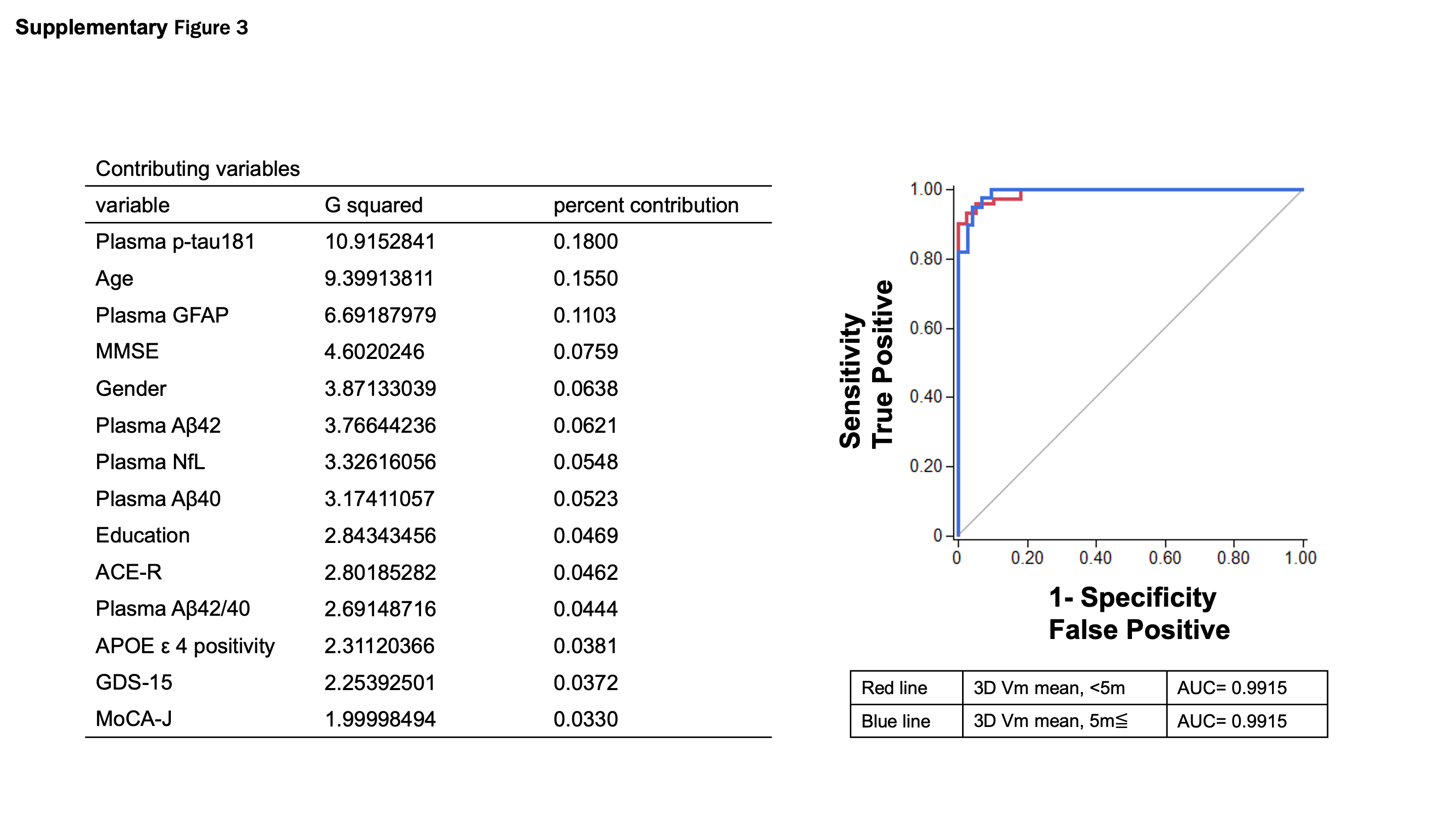

### Figure S4.tiff

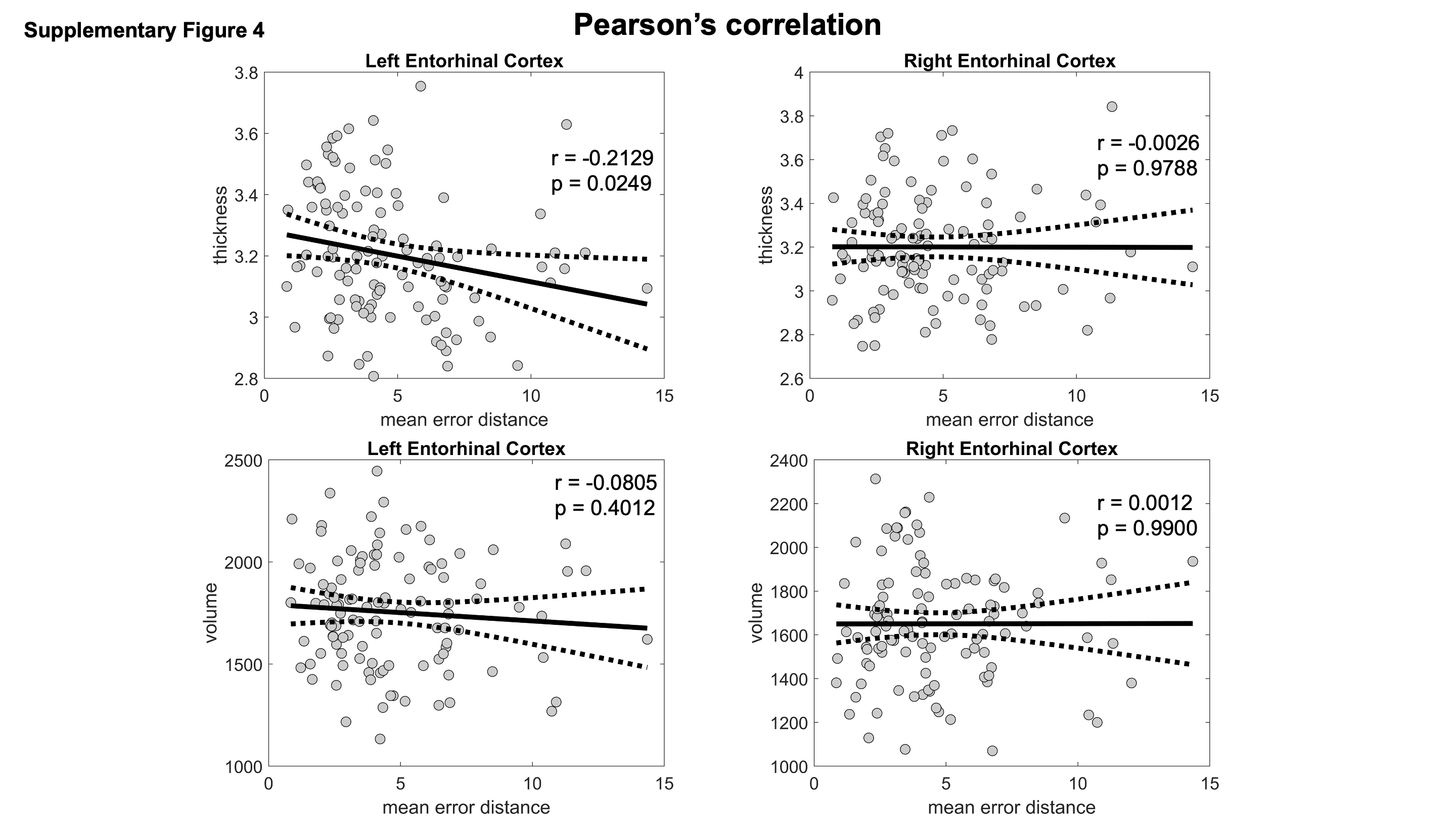

### Figure S5.tiff

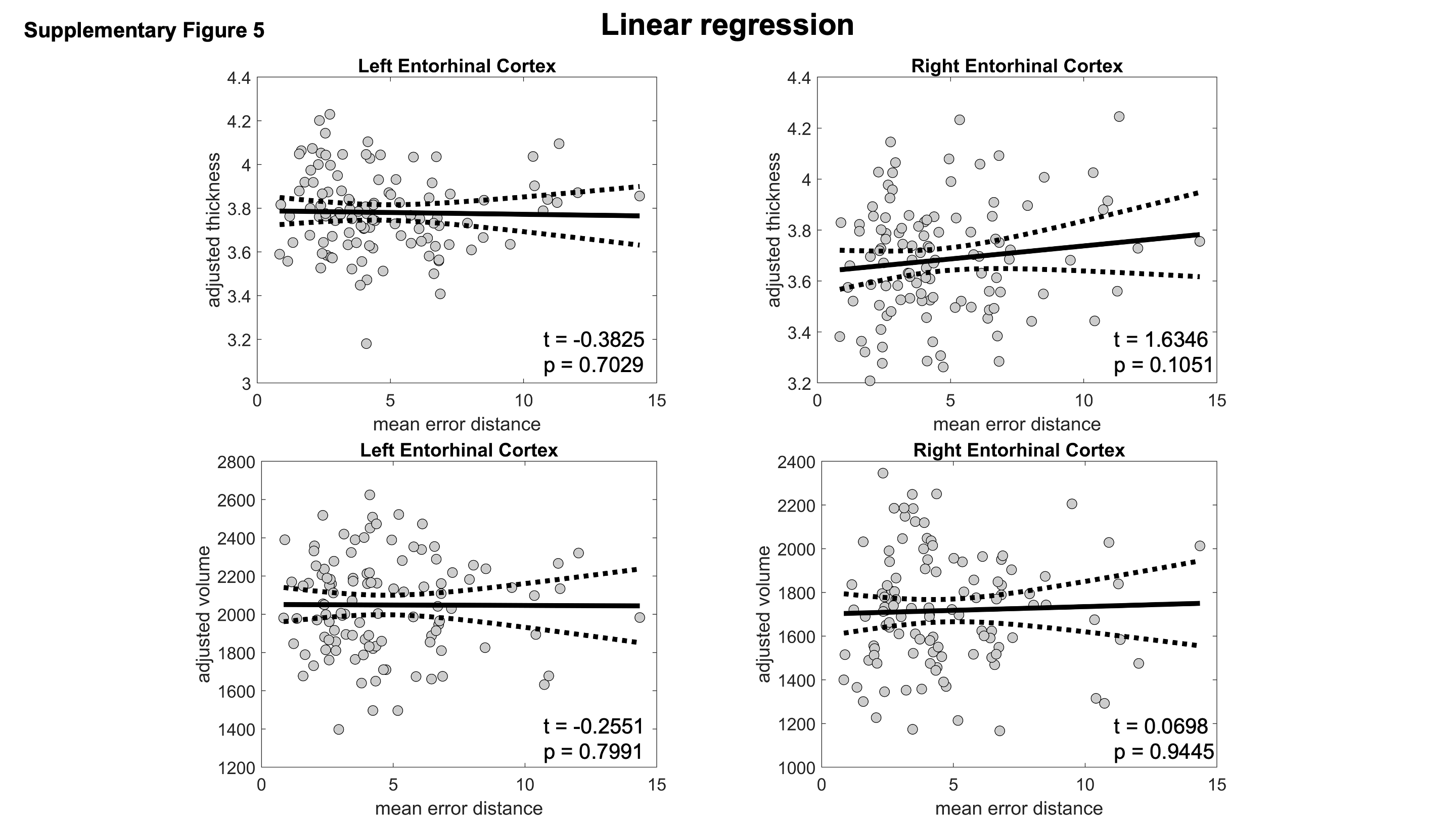
